# Multi-Omics Analysis of Genetic Drivers Linking Aortic Stenosis and Left Ventricular Diastolic Dysfunction in Heart Failure

**DOI:** 10.64898/2026.01.09.26343788

**Authors:** Zeeshan Ahmed, Prithvi Govindareddy, Jayden Mathew, Naveena Yanamala, Partho P. Sengupta

## Abstract

**Background:** Aortic stenosis (AS) and left ventricular diastolic dysfunction (LVDD) often coexist in heart failure (HF), but the mechanisms linking them remain unclear. While AS increases afterload and promotes myocardial stiffening, emerging AI-based evidence suggests LVDD can precede the development of AS or progress simultaneously, indicating shared upstream mechanobiological and inflammatory drivers. This study explores the genetic contributors connecting AS and LVDD to identify early molecular markers and convergent pathways in HF.

**Method:** We analyzed Whole Genome Sequence (WGS) and RNA-seq data of the HF patients, generated using their Peripheral Blood Mononuclear Cells (PBMCs) samples. Overall bioinformatics analysis was divided into two modules, 1) gene variant and annotation analysis, and 2) gene expression and enrichment analysis. We utilized our peer review published and open source WGS and RNA-seq pipelines to process Next-Generation Sequence (NGS) data. Furthermore, we performed bioinformatics and statistical analysis to identify genetic variations, expressions, regulation, enrichments, and disease annotations.

**Results:** We identified genetic markers uniquely associated with AS, LVDD, and shared between them. Furthermore, we report genes with significant expression, and functional variations, and discuss their relationship with other cardiovascular diseases (e.g., *Vascular and Cardiac Stiffness, Aortic Dissection, Left Atrial Enlargement, Left Ventricular Hypertrophy, Outflow Tract Obstructive Defects, Non-Compaction Coronary Artery Disease, Arrhythmia, Congestive Heart Failure, and Hypertrophic, Dilated, and Ischemic Cardiomyopathy*) and non-cardiovascular diseases (non-CVDs) (e.g., *Type 1 Diabetes, Diabetic Nephropathy, Skeletal Anomalies, Rheumatoid Arthritis, Atypical Femoral Fractures, Chronic Kidney Disease, Dehydrated Hereditary Stomatocytosis, Schizophrenia, Varicose Veins, High-Altitude Pulmonary Edema, Periodontitis, and Respiratory Disorder*) including multiple cancer types (e.g., *Breast, Lung, Colorectal, Pancreatic, Hypopharyngeal, Acute Lymphoblastic, and Oral Squamous Cell Carcinomas*) and rare genetic disorders (e.g., *Hypophosphatasia, Multiple Sclerosis, Campomelic Dysplasia, Lymphatic Malformation*). We validated our results through state of science literature, gene-disease annotation databases, and electronic health records.

**Conclusions:** AS and LVDD share both clinical and genomic associations, with overlapping genetic drivers that are enriched in pathways related to inflammation, extracellular matrix remodeling, and vascular stress responses. This work supports the potential of blood-based multi-omics profiling to uncover early, systemic molecular signals of cardiac dysfunction and lays the groundwork for future tissue-specific studies to guide precision diagnosis, risk stratification, and targeted therapeutics in HF.

## 1. Introduction

Heart Failure (HF) is among the most significant causes of morbidity and mortality around the globe [1, 2]. Heart failure with preserved ejection fraction (HFpEF) accounts for nearly half of all HF cases and has traditionally been attributed to myocardial stiffness and impaired diastolic relaxation [3]. Aortic stenosis (AS), long viewed as a valvular disease that causes left ventricular (LV) dysfunction through pressure overload, is now increasingly recognized as being present even in early HFpEF. Large registries, such as the ESC Heart Failure Long-Term Registry, report that approximately 10% of HF patients have aortic valve disease, with AS particularly prevalent in the HFpEF subgroup [4]. Clinical studies further show that mild AS and aortic sclerosis often coexist with subclinical left ventricular diastolic dysfunction (LVDD), suggesting an early and underappreciated connection between these conditions [5]. Furthermore, patients with AS, even after transcatheter valve replacement, show ongoing myocardial stiffening, inflammation, and extracellular-matrix expansion, which influences progression of HFpEF [6].

Recent advances in AI-based predictive models have further challenged the conventional downstream model of AS-induced dysfunction; deep learning algorithms trained on echocardiography and ECG data demonstrate that diastolic dysfunction scores can predict future AS progression, even in patients with anatomically normal valves [7–10]. These findings support the idea that diastolic dysfunction may have a shared pathophysiology with AS. To explain this paradox, we have recently proposed a hypothesis that AS and LVDD arise in parallel from a common mechanoinflammatory environment marked by arterial stiffness, elevated afterload, and disrupted ventriculo–valvular–vascular coupling [11]. This shared biomechanical and inflammatory dynamics provides a unifying framework for understanding the co-evolution of AS and LVDD, particularly in HFpEF.

Despite growing clinical and experimental evidence supporting this model, the underlying genetic architecture that may drive this interconnected pathology remains poorly understood. Evidence from the Framingham Heart Study suggests that cardiovascular disease (CVD) has a complex multifactorial etiology including a genetic component [12]. Multi-omics data, including high-quality sequenced DNA and RNA of transcribed genes, can inform us of a HF patient’s inherent genetic makeup with the most comprehensive view of the genome [13]. DNA-based gene variant detection, when combined with RNA-seq-driven gene expression analysis has the potential to reveal novel genetic markers and stratify HF patient populations based on their disease risk [14, 15].

We conducted a multi-omics investigation using whole genome sequencing (WGS) and RNA sequencing (RNA-seq) of peripheral blood mononuclear cells (PBMCs) from HF patients. This approach allowed us to interrogate both inherited genetic variants and active transcriptional changes, offering a broad view of systemic pathways relevant to cardiac remodeling. PBMCs, although peripheral, reflect immune and inflammatory activity, which plays a central role in both myocardial and valvular pathology. Our analysis integrated variant annotation, gene expression profiling, enrichment analysis, and disease annotation to identify distinct and overlapping genetic signatures associated with AS and LVDD. These findings provide new insight into the molecular mechanisms linking valvular and myocardial dysfunction and support the development of precision diagnostics and targeted therapies for patients at risk of HFpEF.

## 2. Methodology

In this study, we analyzed Whole Genome Sequence (WGS, n=96) and RNA-seq (n=61, with matched WGS samples) data of the HF patients, generated using their PBMCs samples. The WGS cohort was based on 35 female and 60 male cases aged between 29 to 94 years, and one unknown case. However, RNA-seq cohort included 21 female and 40 male cases aged between 45 to 92 years. Furthermore, the RNA-seq dataset was compared against healthy control samples (n=11) without any reported HF diagnosis. Informed consent was obtained from all subjects. All human samples were used in accordance with relevant guidelines and regulations, and all experimental protocols were approved by the Institutional Review Board (IRB) at Rutgers University. All procedures performed in studies involving human participants were in accordance with the ethical standards of the institution and the 1964 Helsinki Declaration and its later amendments or comparable ethical standards. We utilized our peer review published, and open source WGS and RNA-seq pipelines to process Next-Generation Sequence (NGS) data. Furthermore, we performed downstream bioinformatics and statistical analysis to identify target genetic variations, expressions, regulations, enrichments, pathways, and disease annotations. Overall data analysis was divided into two modules, 1) gene variant and annotation, and 2) gene expression and enrichment.

### 2.1. Gene variant and annotation

We processed WGS data (n=96) using JWES [16], which mainly applies Burrows–Wheeler Aligner (BWA) for mapping sequence data against the reference human genome (hg38) [17], and Genome Analysis Toolkit (GATK) for variant discovery and annotation [18]. The outcome of JWES is a Variant Call Format (VCF) file, which we utilized for further downstream analysis, including variant-level quality filtering, classification of coding variations, and identification of functionally relevant alleles associated with AS, LVDD, and both [19]. We utilized Ensembl Variant Effect Predictor (VEP) to predict the effect of variants [20], calculated functional prediction scores using database of Single Nucleotide Polymorphisms (dbSNP) [21], drawn gene-variant-disease relationship with ClinVar [22], and interpreted variant using population allele frequencies from genome Aggregation Database (gnomAD) [23]. We performed functional variant burden analysis and based on the alterations predicted to produce substantial disruption in protein structure and function, we categorized variants into three main categories i.e., high, moderate, and low impact. Furthermore, we utilized annotation databases including Gene Ontology (GO) [24], Kyoto Encyclopedia of Genes and Genomes (KEGG) [25], Reactome Knowledgebase (RK) [26], and Human Phenotype Ontology (HPO) [27] to identify and validate pathogenic variants with potential biological relevance to AS, LVDD, and both. We visualized results, most importantly by generating network diagrams annotating gene-variant and gene-variant-disease relationships.

### 2.2. Gene expression and enrichment

We processed RNA-seq data (n=61) using GVViZ pipeline [28], which mainly applies HISAT (hierarchical indexing for spliced alignment of transcripts) [29] with Bowtie2 [30] to align the sequences against reference hg38, and RSEM (RNA by Expectation Maximization) for the quantification and identification of differentially expressed genes by aligning reads to reference de novo transcriptome assemblies [31]. The major outcome of this pipeline includes quality metrics, isoforms, and gene expressions. Next, we extracted and classified genes relevant to AS, LVDD and both, based on their expression values to perform gene expression, enrichment and annotation analysis. We performed pathway and disease enrichment analysis to identify biological processes and molecular interaction networks overrepresented within the stratified gene sets. Functional enrichment was conducted using the KEGG database to associate cellular functions with stratified gene sets. The enrichment analysis included calculation of False Discovery Rate (FDR) cutoff of 0.05 to control Type I errors and maintain rigorous multiple-testing corrections, enrichment was restricted to pathways with a minimum of two and a maximum of 5,000 genes to eliminate overly broad or narrow biological categories, ensuring results remained relevant to cardiovascular physiology, and redundant terms and overlapping ontologies were filtered to provide a concise, non-redundant functional summary of the biological findings. The top twenty significantly enriched pathways from each dataset were identified and projected to visualize gene enrichment signatures. These visualizations characterized the enrichment ratio, gene count, and statistical significance, highlighting the primary molecular mechanisms and biological themes associated with each HF phenotype.

## 3. Results

We identified through extensive literature review, nine genes known to be associated with AS (*BMP2, TGFB3, ALPL, ITGA5, RUNX2, SPP1, PIEZO1, AREG, SOX9*), nine genes reported for their role in LVDD (*MYBPC3, COL3A1, TGFB2, SOD2, BIRC5, ANKRD1, MYH9, NDRG1, GDF15*), and twenty seven genes shared between both AS and LVDD (*TGFB1, NOS3, IL6, AGT, AGTR1, MPO, NOTCH1, ACE, COL1A1, LGALS3, MMP9, TIMP1, TNF, TNNI3, TNNT2, RHOA, ITGB1, PTK2, FN1, ROCK1, FOXO1, CRP, MMP2, AMOTL2, NPPB, YAP1, TTN*). We report genes with significant expression regulation, functional variations, and their relationship with AS, LVDD, and other CVDs and non-CVDs including multiple cancer types and rare genetic disorders (Table 1 and Table 2).

**Table 1.**
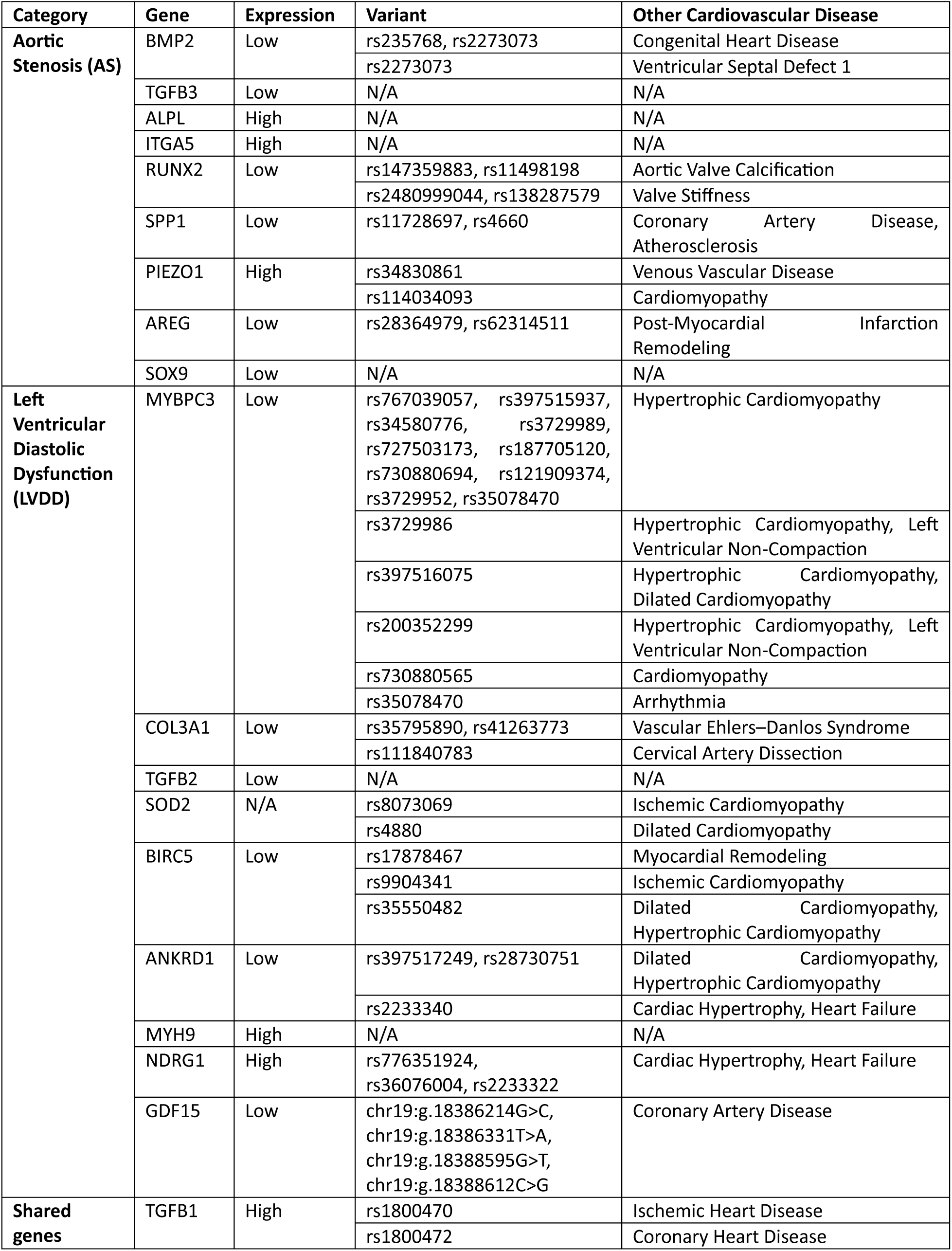
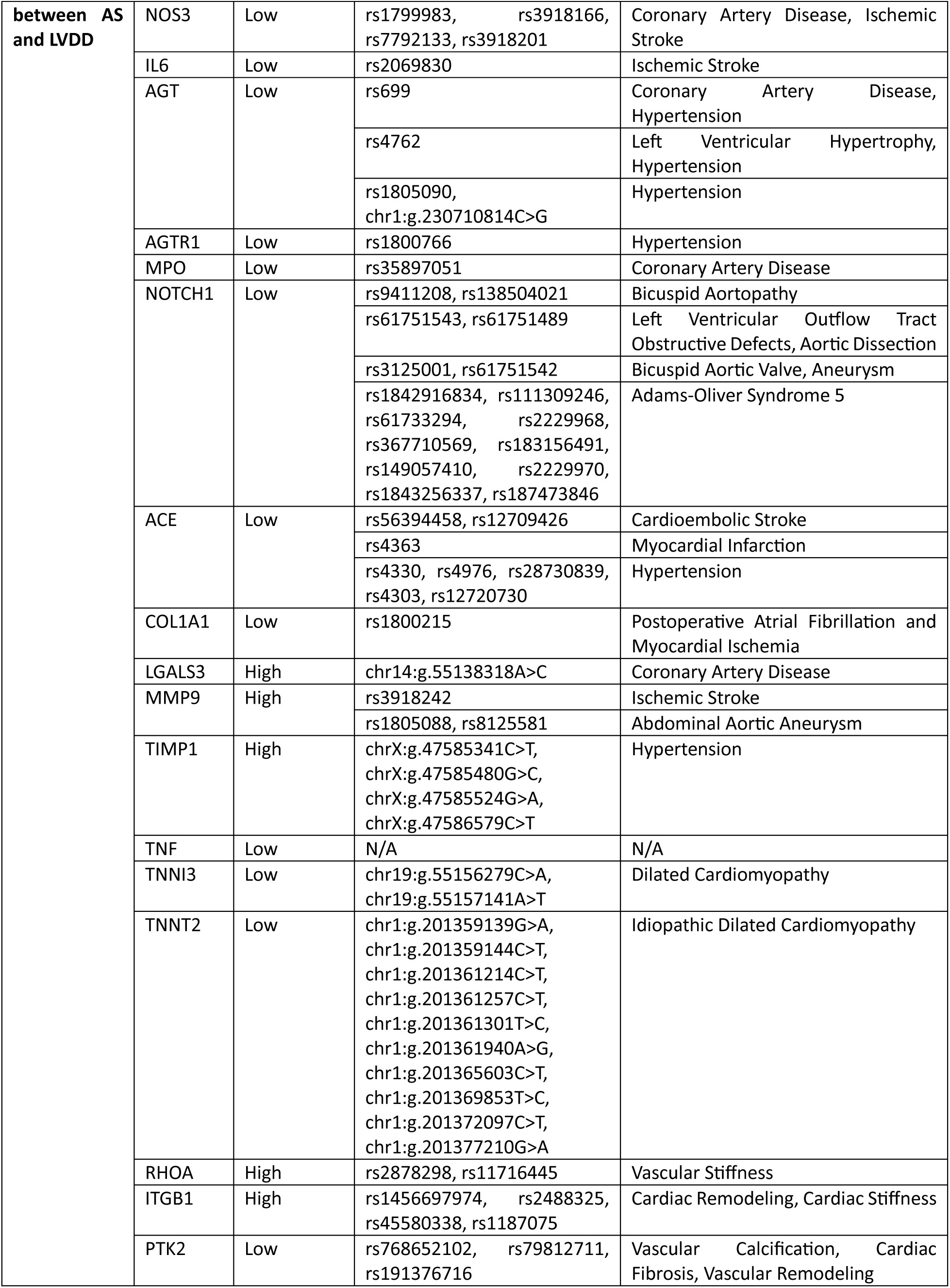
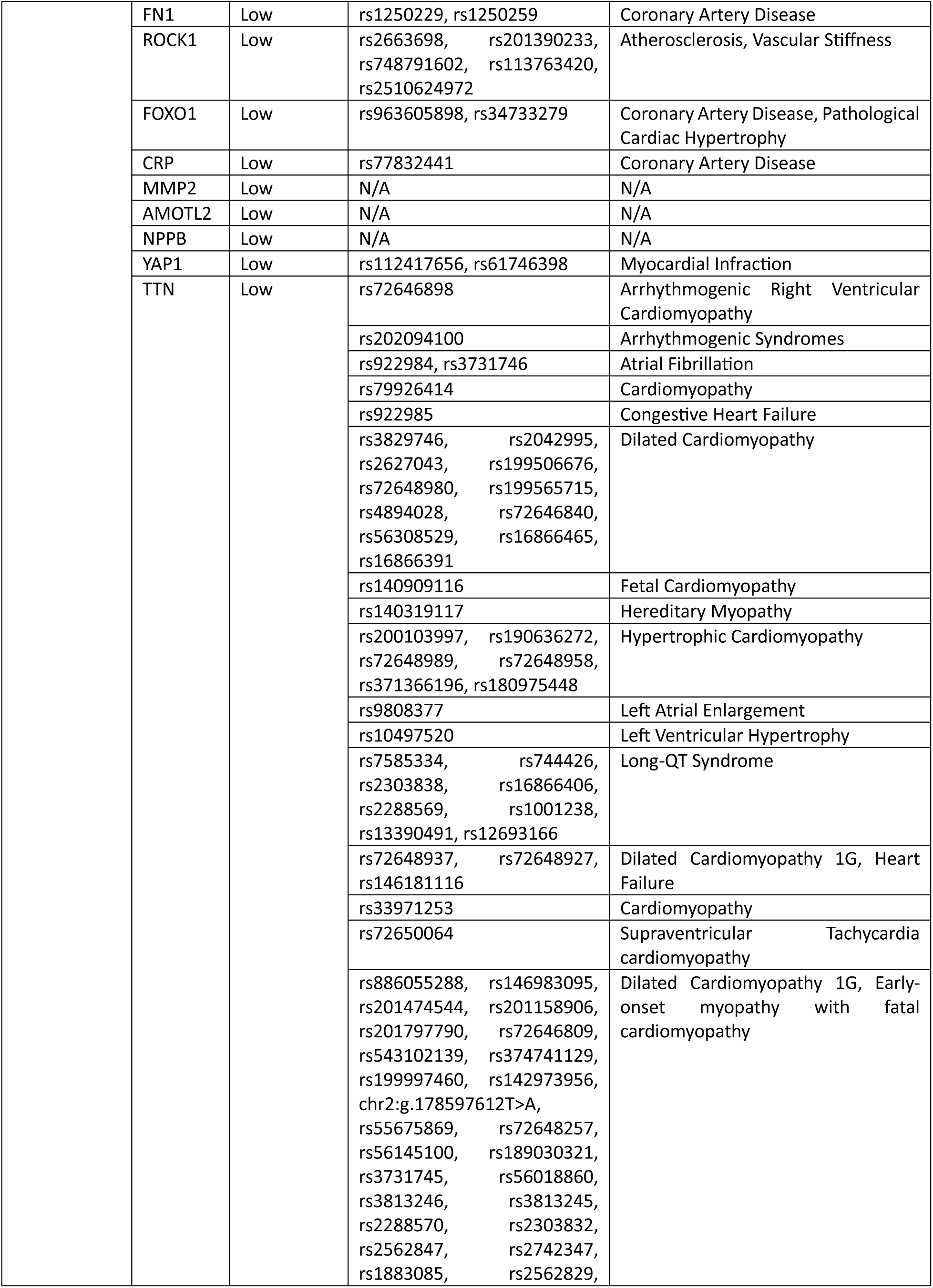
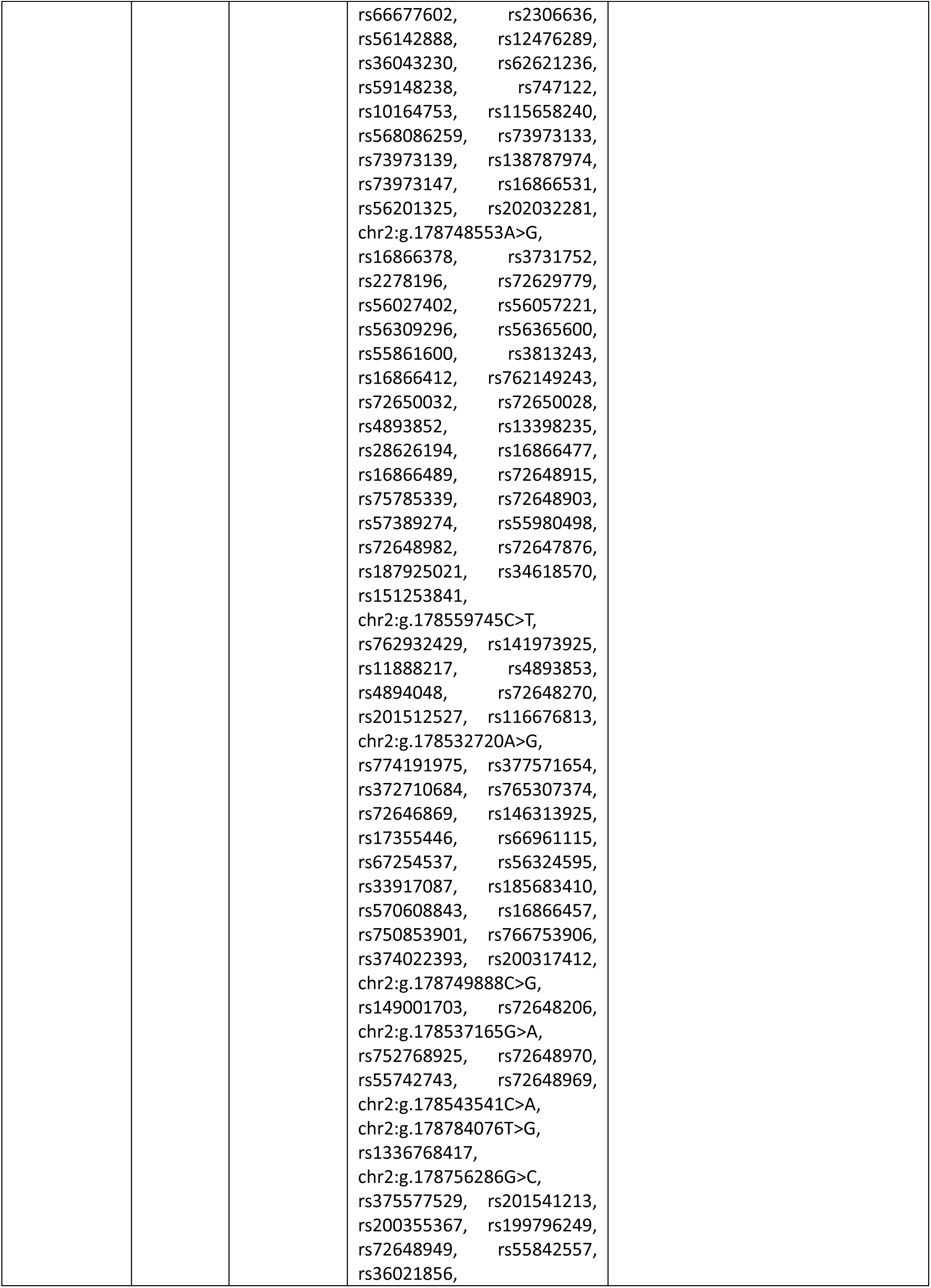
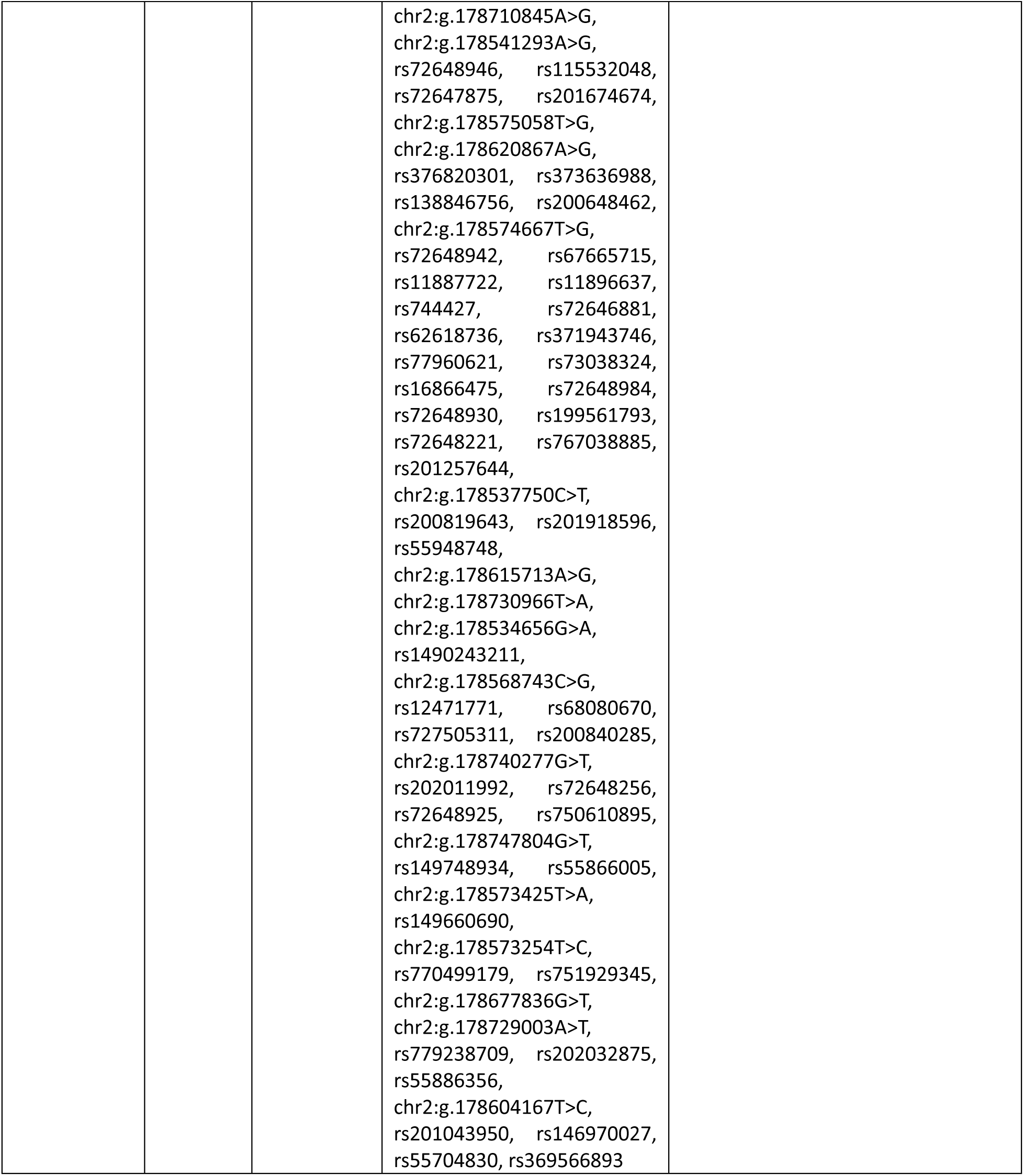
Gene, expression, variants associated with Aortic Stenosis (AS), Left Ventricular Diastolic Dysfunction (LVDD), and shared between AS and LVDD, and other cardiovascular diseases (CVDs).

**Table 2.**
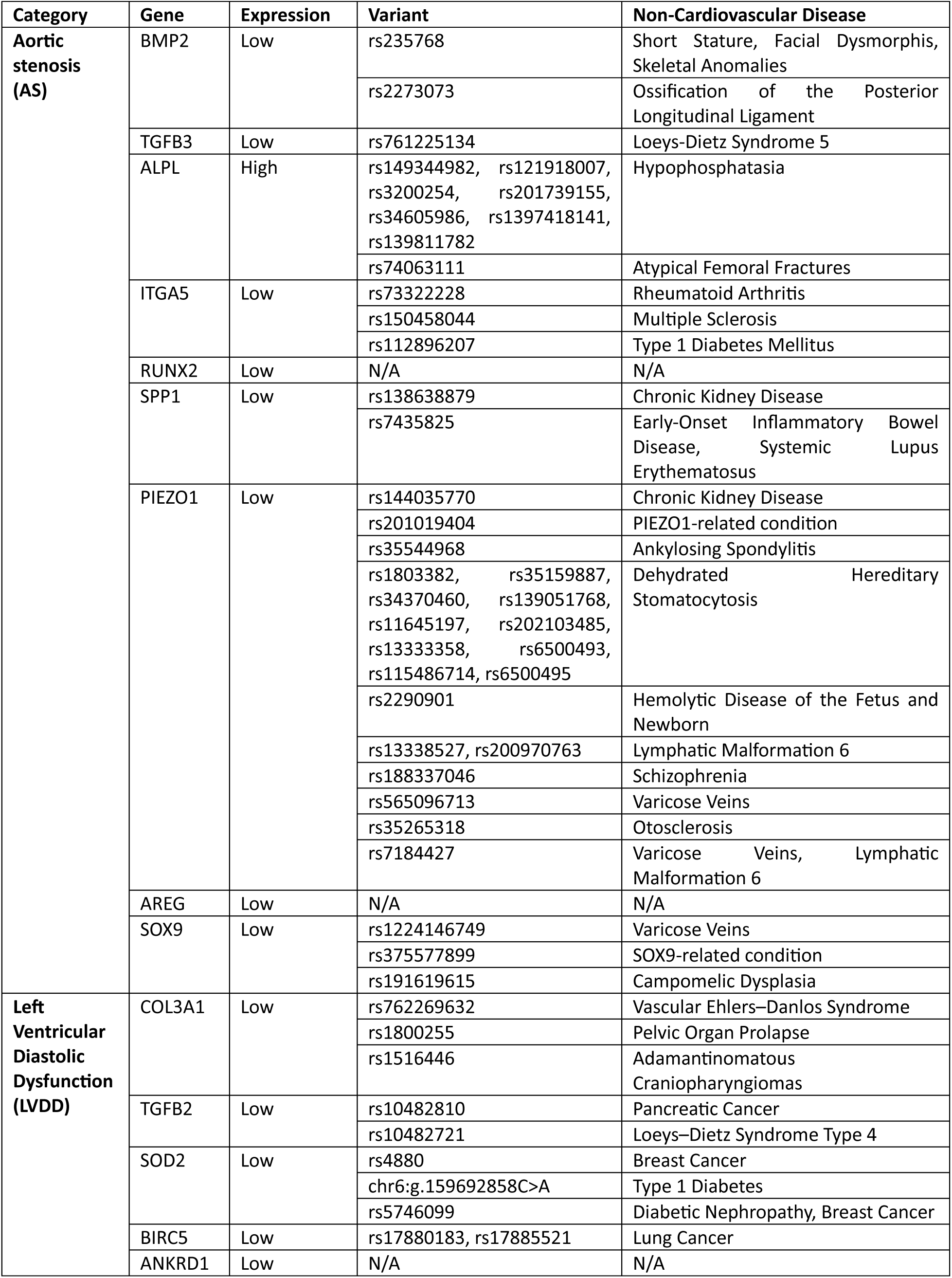
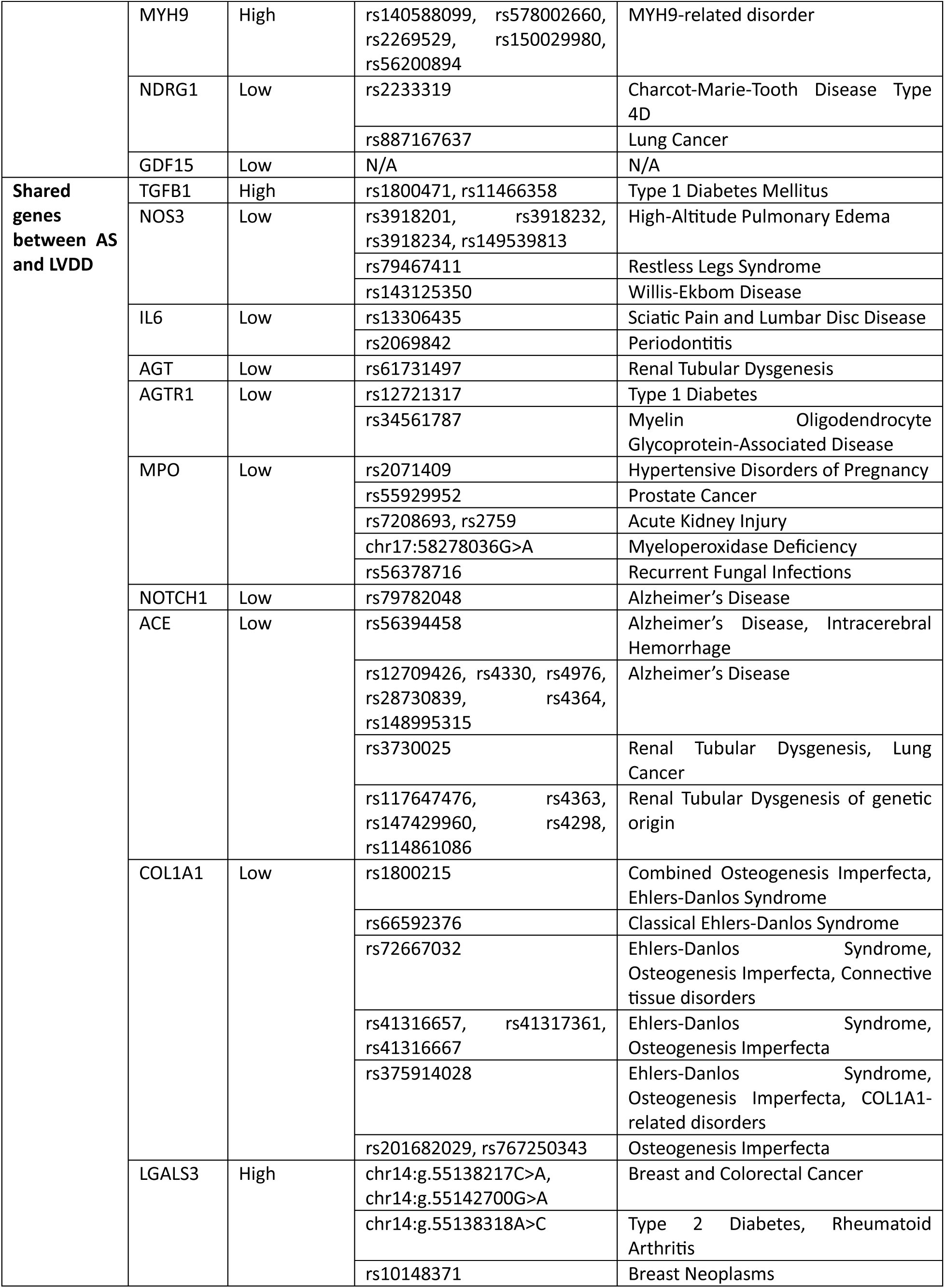
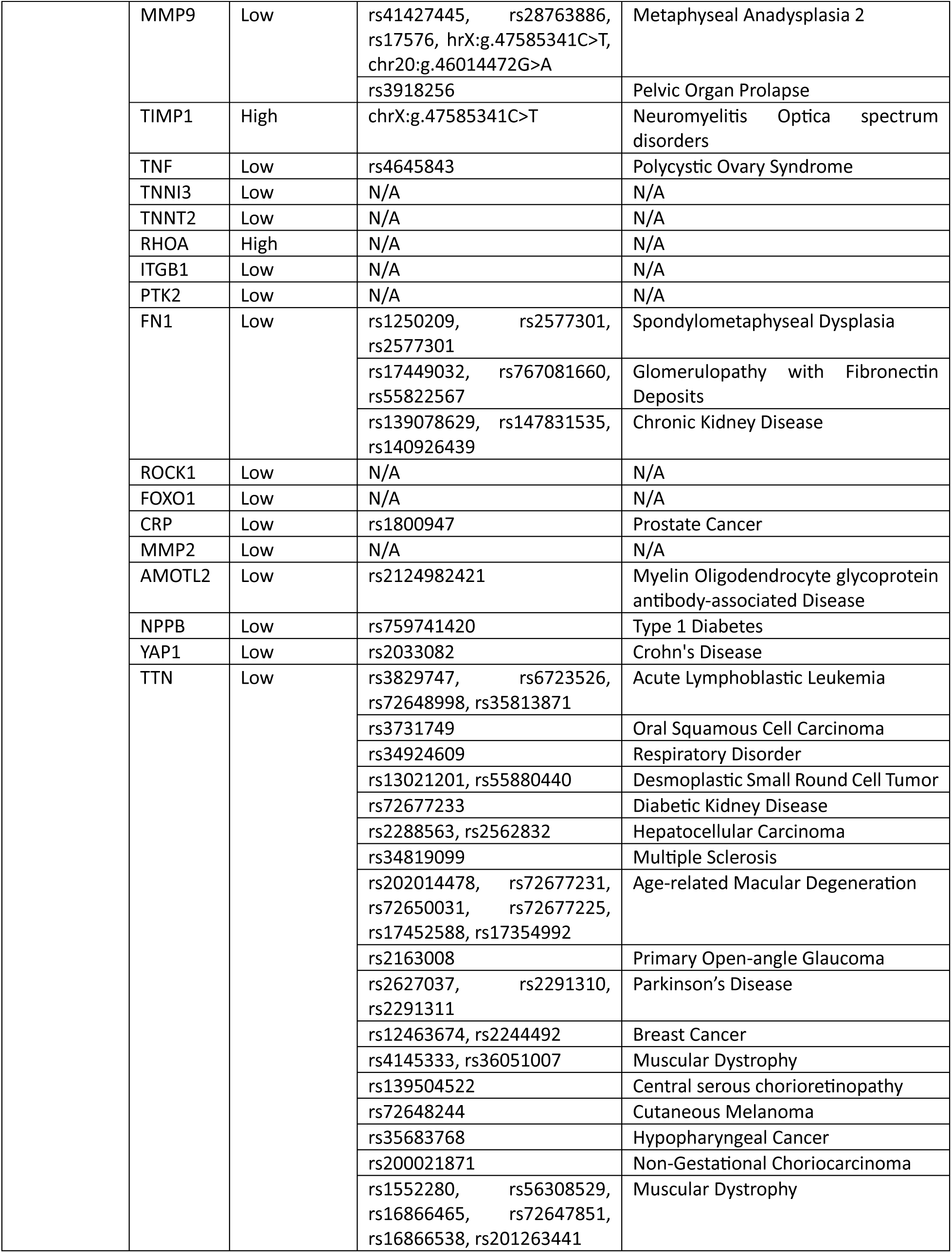
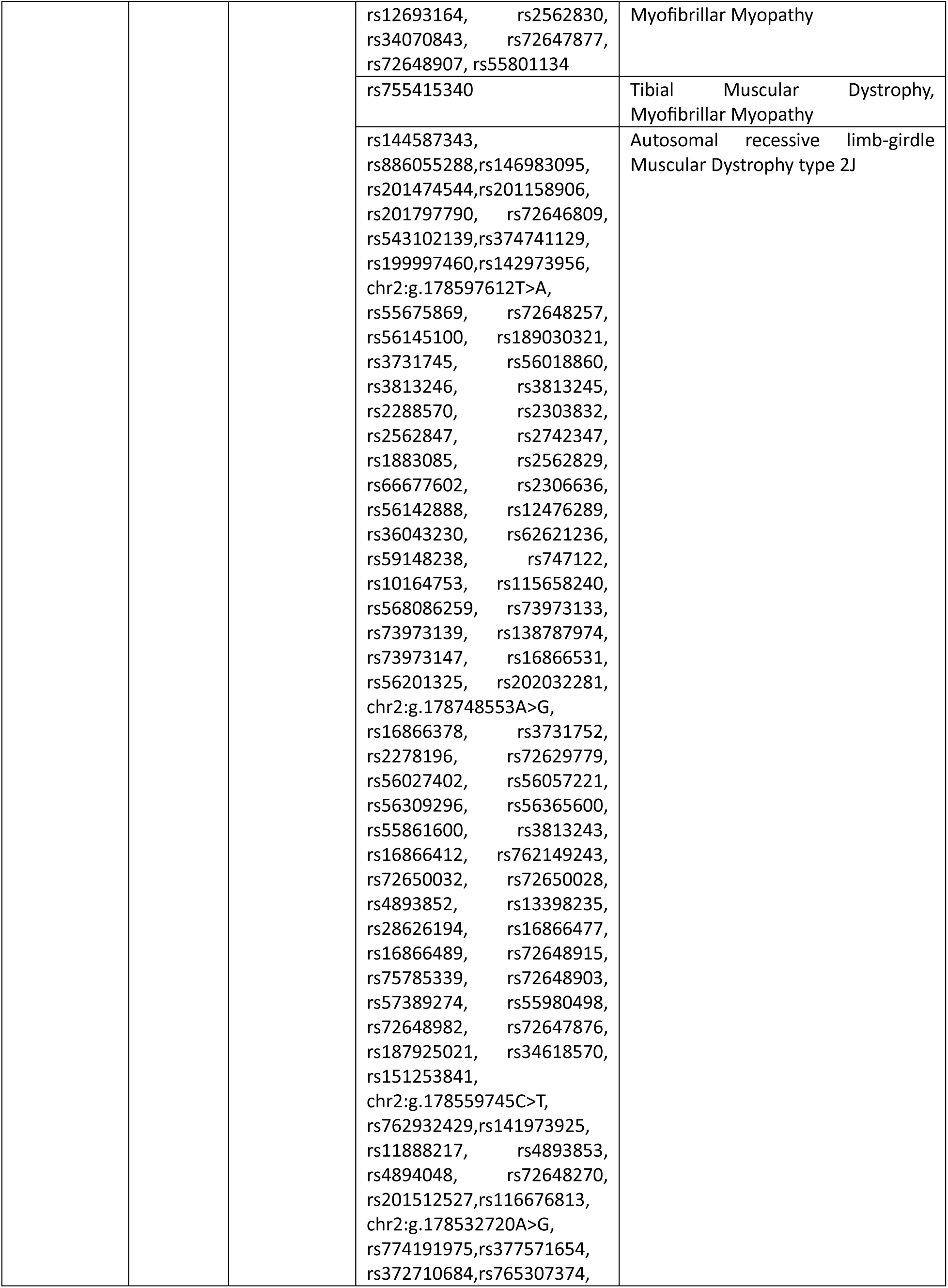
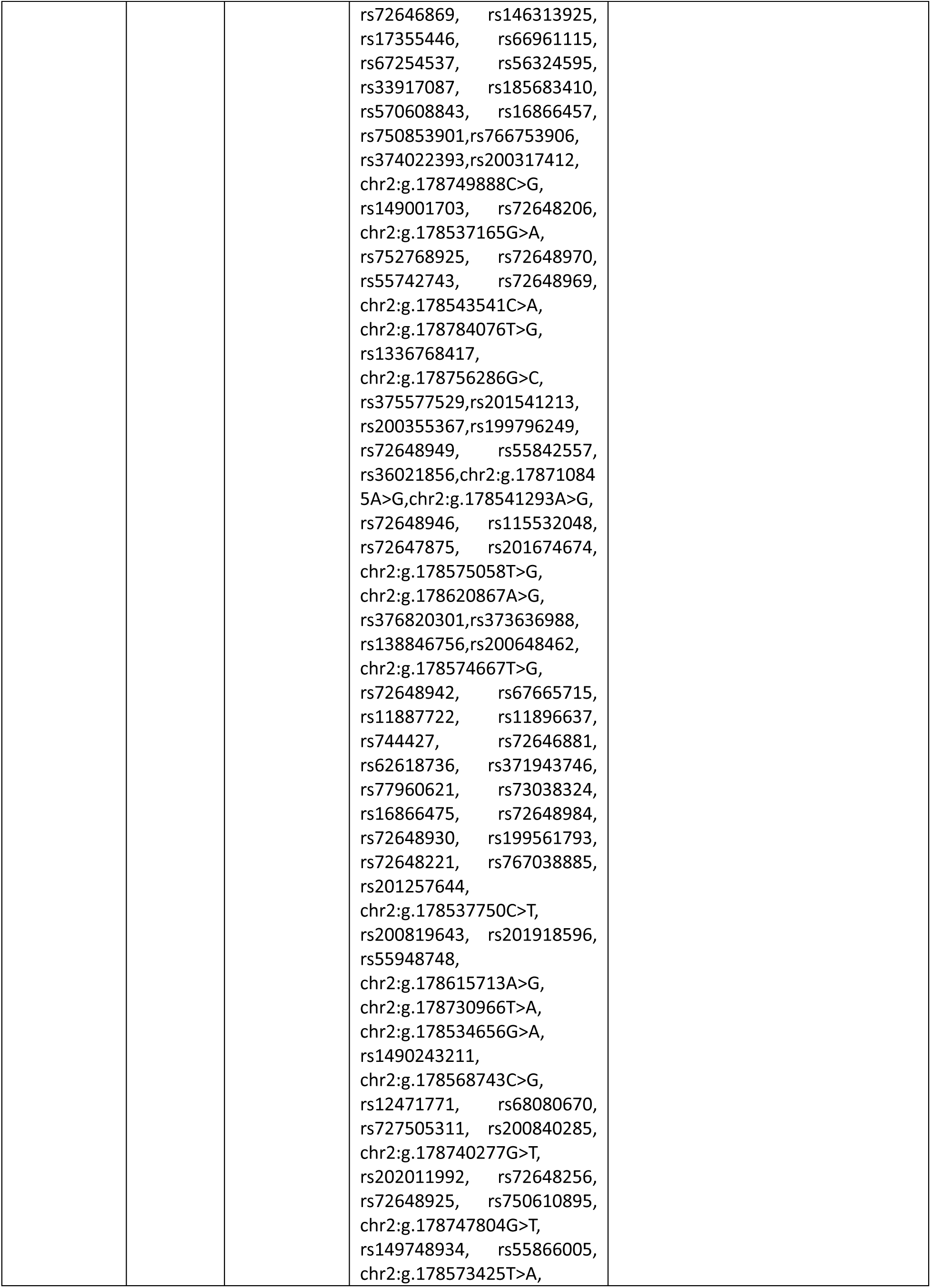
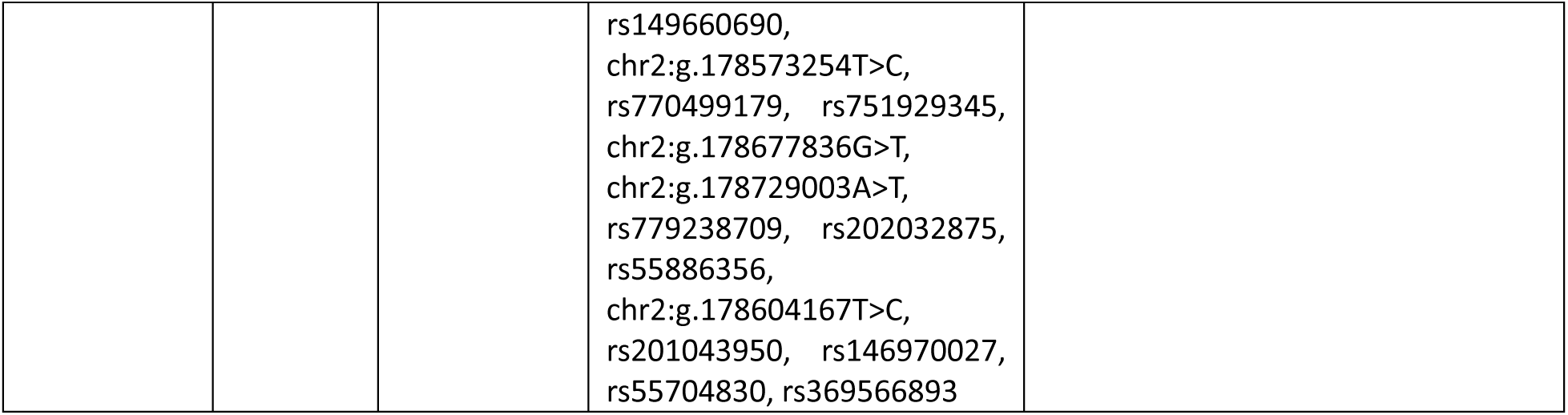
Gene, expression, variants associated with Aortic Stenosis (AS), Left Ventricular Diastolic Dysfunction (LVDD), and shared between AS and LVDD, and non-cardiovascular diseases (non-CVDs).

| Category | Gene | Expression | Variant | Non-Cardiovascular Disease |
| --- | --- | --- | --- | --- |
| Aortic stenosis (AS) | BMP2 | Low | rs235768 | Short Stature, Facial Dysmorphism, Skeletal Anomalies |
|  |  |  | rs2273073 | Ossification of the Posterior Longitudinal Ligament |
|  | TGFB3 | Low | rs761225134 | Loeys-Dietz Syndrome 5 |
|  | ALPL | High | rs149344982, rs121918007, rs3200254, rs201739155, rs34605986, rs1397418141, rs139811782 | Hypophosphatasia |
|  |  |  | rs74063111 | Atypical Femoral Fractures |
|  | ITGA5 | Low | rs73322228 | Rheumatoid Arthritis |
|  |  |  | rs150458044 | Multiple Sclerosis |
|  |  |  | rs112896207 | Type 1 Diabetes Mellitus |
|  | RUNX2 | Low | N/A | N/A |
|  | SPP1 | Low | rs138638879 | Chronic Kidney Disease |
|  |  |  | rs7435825 | Early-Onset Inflammatory Bowel Disease, Systemic Lupus Erythematosus |
|  | PIEZO1 | Low | rs144035770 | Chronic Kidney Disease |
|  |  |  | rs201019404 | PIEZO1-related condition |
|  |  |  | rs35544968 | Ankylosing Spondylitis |
|  |  |  | rs1803382, rs35159887, rs34370460, rs139051768, rs11645197, rs202103485, rs13333358, rs6500493, rs115486714, rs6500495 | Dehydrated Hereditary Stomatocytosis |
|  |  |  | rs2290901 | Hemolytic Disease of the Fetus and Newborn |
|  |  |  | rs13338527, rs200970763 | Lymphatic Malformation 6 |
|  |  |  | rs188337046 | Schizophrenia |
|  |  |  | rs565096713 | Varicose Veins |
|  |  |  | rs35265318 | Otosclerosis |
|  |  |  | rs7184427 | Varicose Veins, Lymphatic Malformation 6 |
|  | AREG | Low | N/A | N/A |
|  | SOX9 | Low | rs1224146749 | Varicose Veins |
|  |  |  | rs375577899 | SOX9-related condition |
|  |  |  | rs191619615 | Campomelic Dysplasia |
| Left Ventricular Diastolic Dysfunction (LVDD) | COL3A1 | Low | rs762269632 | Vascular Ehlers–Danlos Syndrome |
|  |  |  | rs1800255 | Pelvic Organ Prolapse |
|  |  |  | rs1516446 | Adamantinomatous Craniopharyngiomas |
|  | TGFB2 | Low | rs10482810 | Pancreatic Cancer |
|  |  |  | rs10482721 | Loeys–Dietz Syndrome Type 4 |
|  | SOD2 | Low | rs4880 | Breast Cancer |
|  |  |  | chr6:g.159692858C>A | Type 1 Diabetes |
|  |  |  | rs5746099 | Diabetic Nephropathy, Breast Cancer |
|  | BIRC5 | Low | rs17880183, rs17885521 | Lung Cancer |
|  | ANKRD1 | Low | N/A | N/A |
|  | MYH9 | High | rs140588099, rs578002660, rs2269529, rs150029980, rs56200894 | MYH9-related disorder |
|  | NDRG1 | Low | rs2233319 | Charcot-Marie-Tooth Disease Type 4D |
|  |  |  | rs887167637 | Lung Cancer |
|  | GDF15 | Low | N/A | N/A |
| **Shared genes between AS and LVDD** | TGFB1 | High | rs1800471, rs11466358 | Type 1 Diabetes Mellitus |
|  | NOS3 | Low | rs3918201, rs3918232, rs3918234, rs149539813 | High-Altitude Pulmonary Edema |
|  |  |  | rs79467411 | Restless Legs Syndrome |
|  |  |  | rs143125350 | Willis-Ekbom Disease |
|  | IL6 | Low | rs13306435 | Sciatic Pain and Lumbar Disc Disease |
|  |  |  | rs2069842 | Periodontitis |
|  | AGT | Low | rs61731497 | Renal Tubular Dysgenesis |
|  | AGTR1 | Low | rs12721317 | Type 1 Diabetes |
|  |  |  | rs34561787 | Myelin Oligodendrocyte Glycoprotein-Associated Disease |
|  | MPO | Low | rs2071409 | Hypertensive Disorders of Pregnancy |
|  |  |  | rs55929952 | Prostate Cancer |
|  |  |  | rs7208693, rs2759 | Acute Kidney Injury |
|  |  |  | chr17:58278036G>A | Myeloperoxidase Deficiency |
|  |  |  | rs56378716 | Recurrent Fungal Infections |
|  | NOTCH1 | Low | rs79782048 | Alzheimer's Disease |
|  | ACE | Low | rs56394458 | Alzheimer's Disease, Intracerebral Hemorrhage |
|  |  |  | rs12709426, rs4330, rs4976, rs28730839, rs4364, rs148995315 | Alzheimer's Disease |
|  |  |  | rs3730025 | Renal Tubular Dysgenesis, Lung Cancer |
|  |  |  | rs117647476, rs4363, rs147429960, rs4298, rs114861086 | Renal Tubular Dysgenesis of genetic origin |
|  | COL1A1 | Low | rs1800215 | Combined Osteogenesis Imperfecta, Ehlers-Danlos Syndrome |
|  |  |  | rs66592376 | Classical Ehlers-Danlos Syndrome |
|  |  |  | rs72667032 | Ehlers-Danlos Syndrome, Osteogenesis Imperfecta, Connective tissue disorders |
|  |  |  | rs41316657, rs41317361, rs41316667 | Ehlers-Danlos Syndrome, Osteogenesis Imperfecta |
|  |  |  | rs375914028 | Ehlers-Danlos Syndrome, Osteogenesis Imperfecta, COL1A1-related disorders |
|  |  |  | rs201682029, rs767250343 | Osteogenesis Imperfecta |
|  | LGALS3 | High | chr14:g.55138217C>A, chr14:g.55142700G>A | Breast and Colorectal Cancer |
|  |  |  | chr14:g.55138318A>C | Type 2 Diabetes, Rheumatoid Arthritis |
|  |  |  | rs10148371 | Breast Neoplasms |
|  | MMP9 | Low | rs41427445, rs28763886, rs17576, hrX:g.47585341C>T, chr20:g.46014472G>A | Metaphyseal Anadysplasia 2 |
|  |  |  | rs3918256 | Pelvic Organ Prolapse |
|  | TIMP1 | High | chrX:g.47585341C>T | Neuromyelitis Optica spectrum disorders |
|  | TNF | Low | rs4645843 | Polycystic Ovary Syndrome |
|  | TNNI3 | Low | N/A | N/A |
|  | TNNT2 | Low | N/A | N/A |
|  | RHOA | High | N/A | N/A |
|  | ITGB1 | Low | N/A | N/A |
|  | PTK2 | Low | N/A | N/A |
|  | FN1 | Low | rs1250209, rs2577301, rs2577301 | Spondylometaphyseal Dysplasia |
|  |  |  | rs17449032, rs767081660, rs55822567 | Glomerulopathy with Fibronectin Deposits |
|  |  |  | rs139078629, rs147831535, rs140926439 | Chronic Kidney Disease |
|  | ROCK1 | Low | N/A | N/A |
|  | FOXO1 | Low | N/A | N/A |
|  | CRP | Low | rs1800947 | Prostate Cancer |
|  | MMP2 | Low | N/A | N/A |
|  | AMOTL2 | Low | rs2124982421 | Myelin Oligodendrocyte glycoprotein antibody-associated Disease |
|  | NPPB | Low | rs759741420 | Type 1 Diabetes |
|  | YAP1 | Low | rs2033082 | Crohn's Disease |
|  | TTN | Low | rs3829747, rs6723526, rs72648998, rs35813871 | Acute Lymphoblastic Leukemia |
|  |  |  | rs3731749 | Oral Squamous Cell Carcinoma |
|  |  |  | rs34924609 | Respiratory Disorder |
|  |  |  | rs13021201, rs55880440 | Desmoplastic Small Round Cell Tumor |
|  |  |  | rs72677233 | Diabetic Kidney Disease |
|  |  |  | rs2288563, rs2562832 | Hepatocellular Carcinoma |
|  |  |  | rs34819099 | Multiple Sclerosis |
|  |  |  | rs202014478, rs72677231, rs72650031, rs72677225, rs17452588, rs17354992 | Age-related Macular Degeneration |
|  |  |  | rs2163008 | Primary Open-angle Glaucoma |
|  |  |  | rs2627037, rs2291310, rs2291311 | Parkinson's Disease |
|  |  |  | rs12463674, rs2244492 | Breast Cancer |
|  |  |  | rs4145333, rs36051007 | Muscular Dystrophy |
|  |  |  | rs139504522 | Central serous chorioretinopathy |
|  |  |  | rs72648244 | Cutaneous Melanoma |
|  |  |  | rs35683768 | Hypopharyngeal Cancer |
|  |  |  | rs200021871 | Non-Gestational Choriocarcinoma |
|  |  |  | rs1552280, rs56308529, rs16866465, rs72647851, rs16866538, rs201263441 | Muscular Dystrophy |

|  |  |  |  |
| --- | --- | --- | --- |
|  |  | rs12693164, rs2562830,<br>rs34070843, rs72647877,<br>rs72648907, rs55801134 | Myofibrillar Myopathy |
|  |  | rs755415340 | Tibial Muscular Dystrophy,<br>Myofibrillar Myopathy |
|  |  | rs144587343,<br>rs886055288,rs146983095,<br>rs201474544,rs201158906,<br>rs201797790, rs72646809,<br>rs543102139,rs374741129,<br>rs199997460,rs142973956,<br>chr2:g.178597612T>A,<br>rs55675869, rs72648257,<br>rs56145100, rs189030321,<br>rs3731745, rs56018860,<br>rs3813246, rs3813245,<br>rs2288570, rs2303832,<br>rs2562847, rs2742347,<br>rs1883085, rs2562829,<br>rs66677602, rs2306636,<br>rs56142888, rs12476289,<br>rs36043230, rs62621236,<br>rs59148238, rs747122,<br>rs10164753, rs115658240,<br>rs568086259, rs73973133,<br>rs73973139, rs138787974,<br>rs73973147, rs16866531,<br>rs56201325, rs202032281,<br>chr2:g.178748553A>G,<br>rs16866378, rs3731752,<br>rs2278196, rs72629779,<br>rs56027402, rs56057221,<br>rs56309296, rs56365600,<br>rs55861600, rs3813243,<br>rs16866412, rs762149243,<br>rs72650032, rs72650028,<br>rs4893852, rs13398235,<br>rs28626194, rs16866477,<br>rs16866489, rs72648915,<br>rs75785339, rs72648903,<br>rs57389274, rs55980498,<br>rs72648982, rs72647876,<br>rs187925021, rs34618570,<br>rs151253841,<br>chr2:g.178559745C>T,<br>rs762932429,rs141973925,<br>rs11888217, rs4893853,<br>rs4894048, rs72648270,<br>rs201512527,rs116676813,<br>chr2:g.178532720A>G,<br>rs774191975,rs377571654,<br>rs372710684,rs765307374, | Autosomal recessive limb-girdle<br>Muscular Dystrophy type 2J |

|  |  |  |  |
| --- | --- | --- | --- |
|  |  |  | rs72646869, rs146313925,<br>rs17355446, rs66961115,<br>rs67254537, rs56324595,<br>rs33917087, rs185683410,<br>rs570608843, rs16866457,<br>rs750853901,rs766753906,<br>rs374022393,rs200317412,<br>chr2:g.178749888C>G,<br>rs149001703, rs72648206,<br>chr2:g.178537165G>A,<br>rs752768925, rs72648970,<br>rs55742743, rs72648969,<br>chr2:g.178543541C>A,<br>chr2:g.178784076T>G,<br>rs1336768417,<br>chr2:g.178756286G>C,<br>rs375577529,rs201541213,<br>rs200355367,rs199796249,<br>rs72648949, rs55842557,<br>rs36021856,chr2:g.17871084<br>5A>G,chr2:g.178541293A>G,<br>rs72648946, rs115532048,<br>rs72647875, rs201674674,<br>chr2:g.178575058T>G,<br>chr2:g.178620867A>G,<br>rs376820301,rs373636988,<br>rs138846756,rs200648462,<br>chr2:g.178574667T>G,<br>rs72648942, rs67665715,<br>rs11887722, rs11896637,<br>rs744427, rs72646881,<br>rs62618736, rs371943746,<br>rs77960621, rs73038324,<br>rs16866475, rs72648984,<br>rs72648930, rs199561793,<br>rs72648221, rs767038885,<br>rs201257644,<br>chr2:g.178537750C>T,<br>rs200819643, rs201918596,<br>rs55948748,<br>chr2:g.178615713A>G,<br>chr2:g.178730966T>A,<br>chr2:g.178534656G>A,<br>rs1490243211,<br>chr2:g.178568743C>G,<br>rs12471771, rs68080670,<br>rs727505311, rs200840285,<br>chr2:g.178740277G>T,<br>rs202011992, rs72648256,<br>rs72648925, rs750610895,<br>chr2:g.178747804G>T,<br>rs149748934, rs55866005,<br>chr2:g.178573425T>A, rs149660690,<br>chr2:g.178573254T>C,<br>rs770499179, rs751929345,<br>chr2:g.178677836G>T,<br>chr2:g.178729003A>T,<br>rs779238709, rs202032875,<br>rs55886356,<br>chr2:g.178604167T>C,<br>rs201043950, rs146970027,<br>rs55704830, rs369566893 |

### 3.1. Gene variant profiling and annotation

The WGS data analysis, uncovered functional variants from genes associated with AS, LVDD, and may have convergent role in AS and LVDD. Furthermore, we investigated their relationships with other CVDs.

#### AS associated gene-variant profiling

CVD associations for AS-related genes were obtained through gene-variant-disease network analysis (Figures 1A). Variant profiling disclosed variants for five AS genes (Figures 2A, Table 1). Only two likely moderate variants were identified each for *AREG*, *PIEZO1*, and *SPP1* with one high impact variant for *SPP1*. *BMP2* and *RUNX2, however,* had three and four moderate variants identified respectively. Associations between these five genes and CVDs were limited. *BMP2* showed three functional variants (*rs235768, rs2273073, rs2273073*), observed across eighty-four participants, associated with congenital heart disease-related phenotypes, including ventricular septal defects. *AREG* exhibited two functional variants (*rs28364979, rs62314511*) in seventy-two participants, which were related to post-myocardial infarction remodeling. *PIEZO1* displayed two functional variants in eighty-four participants linked to venous vascular disease (*rs34830861*) and cardiomyopathy (*rs114034093*). *SPP1* exhibited two protein-altering variants (*rs11728697, rs4660*) in sixty-six participants, strongly impactful for coronary artery disease and atherosclerosis. *RUNX2* showed four functional variants in fifty-four participants, associated with aortic valve calcification (*rs147359883, rs11498198*) and valve stiffness (*rs2480999044, rs138287579*).

**Figure 1.**
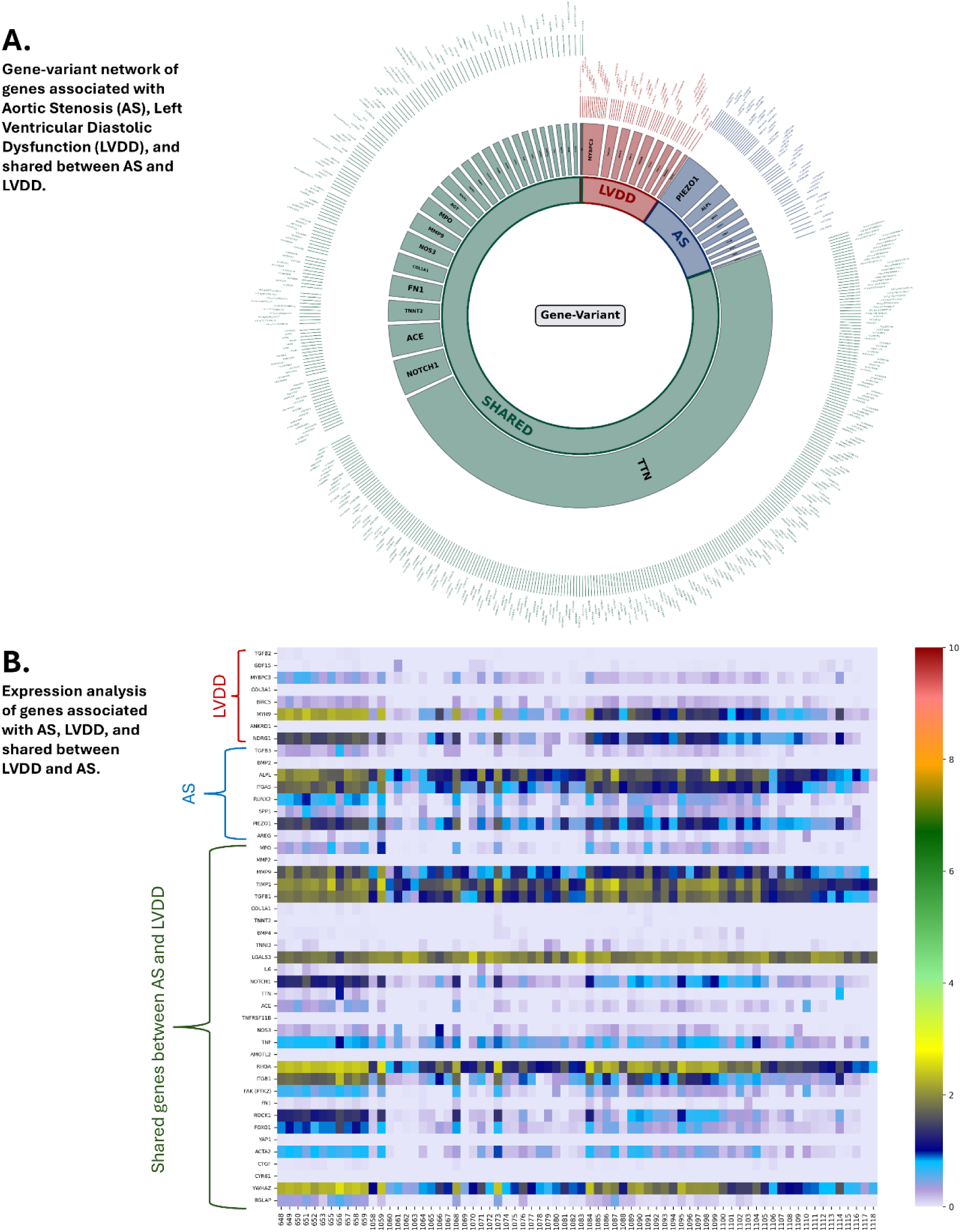
(A) Gene-variant network of genes associated with Aortic Stenosis (AS), Left Ventricular Diastolic Dysfunction (LVDD), and shared between AS and LVDD. **(B)** Expression analysis of genes associated with AS, LVDD, and shared between LVDD and AS.

**Figure 2.**
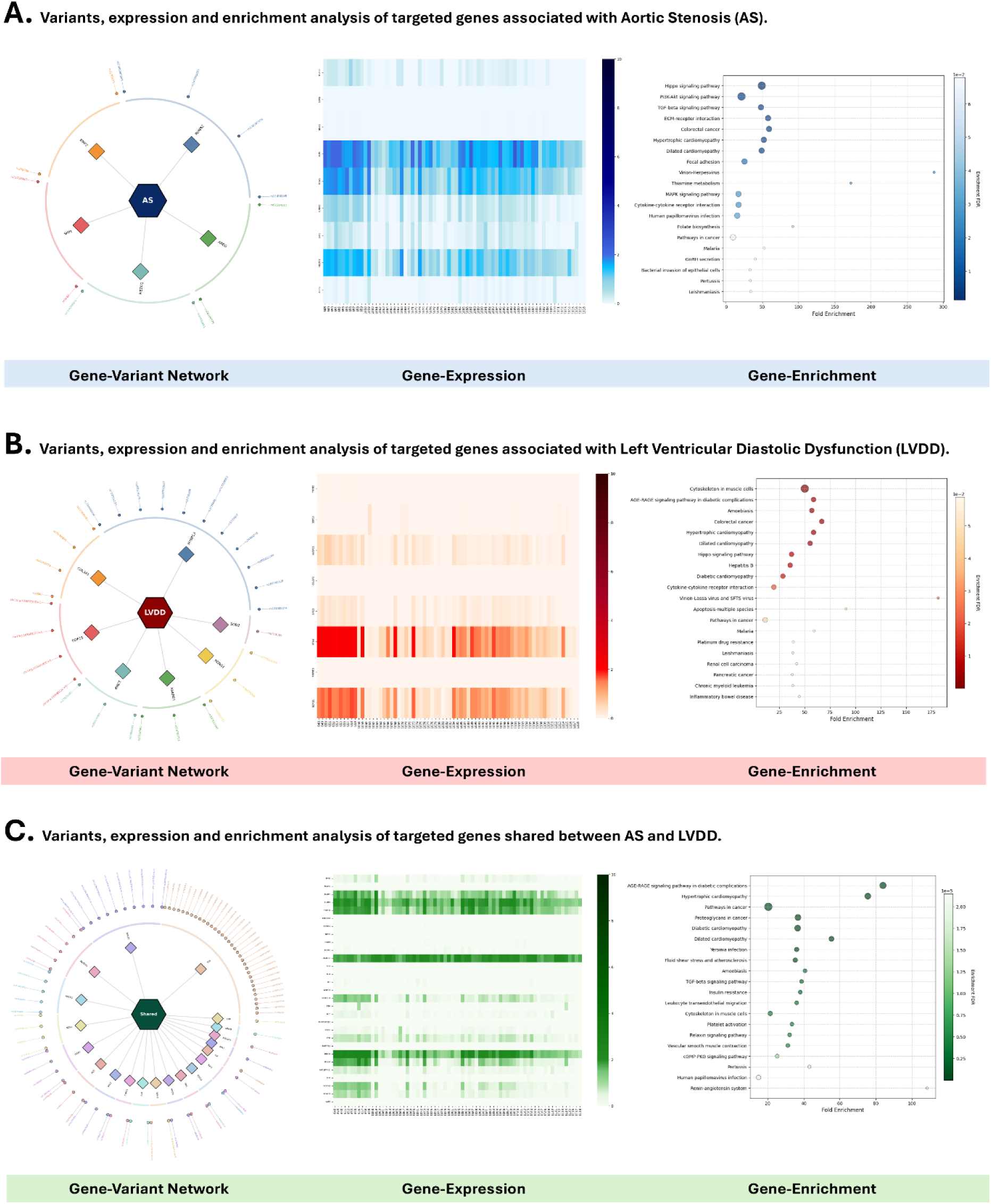
(A) Variants, expression and enrichment analysis of targeted genes associated with Aortic Stenosis (AS). **(B)** Variants, expression and enrichment analysis of targeted genes associated with Left Ventricular Diastolic Dysfunction (LVDD). **(C)** Variants, expression and enrichment analysis of targeted genes shared between AS and LVDD.

#### LVDD associated gene-variant profiling

CVD associations for LVDD-related genes were derived from gene-variant-disease network analysis (Figure 1A). Variant profiling disclosed variants for seven LVDD genes (Figures 2B, Table 1). Fifteen variants were identified for *MYBPC3* with two high impact variants. Three moderate variants were identified for *COL3A1*, *BIRC5*, *ANKRD1,* and *NDRG1* with one *BIRC5* variant as highly pathogenic. Four moderate variants were found for *GDF15* and two for *SOD2.* Among the curated LVDD-associated genes, *MYBPC3* showed fifteen distinct variants in forty-eight participants. These variants were found to be associated with cardiomyopathy-related phenotypes, including two protein-altering variants being highly pathogenic for hypertrophic cardiomyopathy (*rs767039057, rs397515937, rs34580776, rs3729989, rs727503173, rs187705120, rs730880694, rs121909374, rs3729952, rs35078470, rs3729986, rs397516075, rs200352299*), dilated cardiomyopathy (*rs397516075, rs730880565*), left ventricular non-compaction (*rs3729986, rs200352299*), and Arrhythmia (*rs35078470*). *COL3A1* displayed three functional variants across all participants, linked to Vascular Ehlers–Danlos Syndrome (*rs35795890, rs41263773*) and Cervical Artery Dissection (*rs111840783*). *SOD2* harbored two functional variants in seventy-three participants, associations included ischemic cardiomyopathy (*rs8073069*) and dilated cardiomyopathy (*rs4880*). *GDF15* exhibited four functional variants (*chr19:g.18386214G>C, chr19:g.18386331T>A, chr19:g.18388595G>T, chr19:g.18388612C>G*) in eighty-five participants, related to coronary artery disease. *BIRC5* showed three functional variants in seventy-five participants, and its CVD associations included one high impact protein altering variant for myocardial remodeling (rs17878467), ischemic cardiomyopathy (rs9904341), dilated cardiomyopathy, and hypertrophic cardiomyopathy (rs35550482). *ANKRD1* showed three functional variants in sixty-three participants, connected to dilated cardiomyopathy and hypertrophic cardiomyopathy (*rs397517249, rs28730751*), and cardiac hypertrophy and HF (*rs2233340*). *NDRG1* showed three functional variants in eighty-one participants, reported for cardiac hypertrophy and HF (*rs776351924, rs36076004, rs2233322*).

#### Gene-variant profiling of genes shared between AS and LVDD

We identified twenty-seven genes with shared biology in AS and LVDD (Figures 1A, and Figure 2A and B). However, variant profiling disclosed variants for twenty-four genes associated with other CVDs (Figure 2C, and Table 1). These genes include *TGFB1, NOS3, IL6, AGT, AGTR1, MPO, NOTCH1, ACE, COL1A1, LGALS3, MMP9, TIMP1, TNF, TNNI3, TNNT2, RHOA, ITGB1, PTK2, FN1, ROCK1, FOXO1, CRP, YAP1, TTN*.

We found two functional variants in *TGFB1* across seventy-three participants, which were associated with ischemic heart disease (*rs1800470*) and coronary heart disease (*rs1800472*). *NOS3* showed four functional variants (*rs1799983, rs3918166, rs7792133, rs3918201*) across fifty-seven participants and linked to coronary artery disease and ischemic stroke. *AGT* displayed four functional variants within seventy-four participants, related to coronary artery disease and hypertension (*rs699*), left ventricular hypertrophy (*rs4762*) and hypertension (*rs1805090, chr1:g.230710814C>G*). *NOTCH1* showed sixteen functional variants observed in eighty-eight participants, related to bicuspid aortopathy (*rs9411208, rs138504021*), left ventricular outflow tract obstructive defects, aortic dissection (*rs61751543, rs61751489*), bicuspid aortic valve, aneurysm (*rs3125001, rs61751542*), and Adams-Oliver syndrome 5 (*rs1842916834, rs111309246, rs61733294, rs2229968, rs367710569, rs183156491, rs149057410, rs2229970, rs1843256337, rs187473846*). *ACE* revealed eight functional variants in ninety-six participants with one high impact variant associated with hypertension (*rs4330, rs4976, rs28730839, rs4303, rs12720730*), cardioembolic stroke (*rs56394458, rs12709426*), and myocardial infarction (*rs4363*). *MMP9* expressed three functional variants identified in ninety-six participants, linked with ischemic stroke (*rs3918242*) and abdominal aortic aneurysm (*rs1805088, rs8125581*). *TIMP1* showed four functional variants observed in fifty-four participants, associated with hypertension (*chrX:g.47585341C>T, chrX:g.47585480G>C, chrX:g.47585524G>A, chrX:g.47586579C>T*). *TNNT2* expressed ten functional variants (*chr1:g.201359139G>A, chr1:g.201359144C>T, chr1:g.201361214C>T, chr1:g.201361257C>T, chr1:g.201361301T>C, chr1:g.201361940A>G, chr1:g.201365603C>T, chr1:g.201369853T>C, chr1:g.201372097C>T, chr1:g.201377210G>A*) within twenty-four participants, which were linked to idiopathic dilated cardiomyopathy. *ITGB1* showed four functional variants (*rs1456697974, rs2488325, rs45580338, rs1187075*) identified in seventy-one participants, with one highly damaging variant linked to cardiac remodeling and cardiac stiffness. *PTK2* expressed three functional variants (*rs768652102, rs79812711, rs191376716*) detected in forty-eight participants, associated with vascular calcification, cardiac fibrosis, and vascular remodeling. *ROCK1* showed five functional variants (*rs2663698, rs201390233, rs748791602, rs113763420, rs2510624972*) observed in fifty-seven participants, with one high impact variant associated with atherosclerosis and vascular stiffness. Few genes reported just two functional variants associated with similar CVDs, which include *TNNI3* (*chr19:g.55156279C>A and chr19:g.55157141A>T;* dilated cardiomyopathy) identified in forty-four participants, *RHOA* (*rs2878298 and rs11716445;* vascular stiffness) observed in sixty-two participants, *FN1* (*rs1250229 and rs1250259;* coronary artery disease) present in eighty-six participants, *FOXO1* (*rs963605898* and *rs34733279*; coronary artery disease and pathological cardiac hypertrophy) identified in sixty-nine participants, and *YAP1* (*rs112417656* and *rs61746398*; myocardial infraction) observed in fifty-two participants with one high impact variant.

In addition to these, a single CVD related functional variant was exhibited by the following genes; *IL6* (*rs2069830l;* ischemic stroke) observed across sixty-five participants, a high impact variant for *AGTR1* (*rs1800766;* hypertension) within twenty-seven participants and *MPO* (*rs35897051l* coronary artery disease) within fifty-one participants, *COL1A1 (rs1800215;* postoperative atrial fibrillation and myocardial ischemia) detected in fifteen participants, *LGALS3* (*chr14:g.55138318A>C;* coronary artery disease), and *CRP* (*rs77832441;* coronary artery disease) detected in fourteen participants.

We identified most of the functional variants (n= 276, Table 1) in *TTN* among ninety-one participants, which were associated with variable AS, LVDD, and many other CVDs including arrhythmogenic right ventricular cardiomyopathy, arrhythmogenic syndromes, atrial fibrillation, cardiomyopathy, congestive HF, dilated cardiomyopathy, fetal cardiomyopathy, hereditary myopathy, hypertrophic cardiomyopathy, left atrial enlargement, left ventricular hypertrophy, long-QT syndrome, dilated cardiomyopathy 1G, HF, cardiomyopathy, supraventricular tachycardia cardiomyopathy, and, early-onset myopathy with fatal cardiomyopathy. Detailed classification is provided in Table 1 (Figure 3A and B).

**Figure 3.**
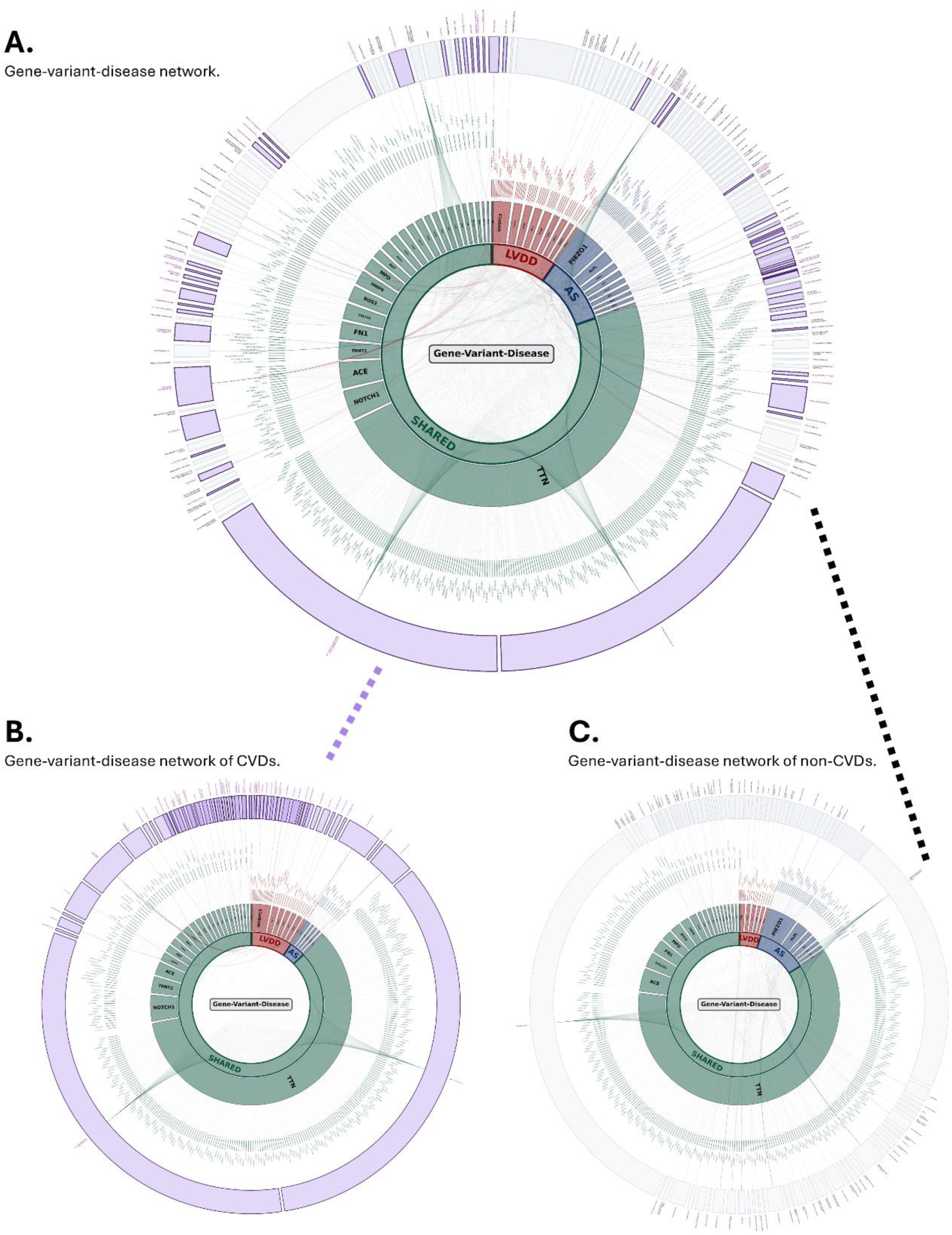
(A) Gene-variant-disease network of genes associated with Aortic Stenosis (AS), Left Ventricular Diastolic Dysfunction (LVDD), and shared between AS and LVDD. **(B)** Gene-variant-disease network of genes associated with other cardiovascular diseases (CVDs). **(C)** Gene-variant-disease network of genes associated with non-cardiovascular diseases (non-CVDs).

### 3.2. Gene expression and enrichment

Integrating In-house developed and published bioinformatics pipelines with systematic expression stratification and pathway enrichment, we ensured the precise identification of transcriptionally active genes that drive the biological convergence between AS and LVDD phenotypes leading to HF. Transcriptomic profiling across the case group (IDs 1058–1118) and the control group (IDs 648–659) was performed in two steps. First, gene expression analysis of all genes known to be associated with AS, LVDD, and have a strong overlapping between both AS and LVDD (Figure 1B, and Figure 2A, B, and C). Second, gene enrichment analysis of all genes known to be associated with AS, LVDD, and strongly intersect between AS and LVDD (Figure 2A, B, and C).

#### AS-associated gene expression analysis

AS-specific gene expression identified heatmap-based expression for all nine markers with major association to vascular mineralization and cardiac remodeling (Figure 2A). *ALPL*, *ITGA5*, and *PIEZO1* showed comparably higher expression in the control groups with significant variations observed in case subsets. *ALPL* was the highly expressed gene in this subset, with significantly high intensity in the control group. Gene expression profiling showed variable intensity in *ALPL* expression in case samples (Figure 2A). CVD found coronary calcification [32] and mineralization-related disorders [33]. *ITGA5* also exhibited a high-intensity profile that was very strong in the control group in comparison to specific elderly case subjects. Expression analysis indicated high *ITGA5* expression across the control group (Figure 2A). CVD associations involved cell-matrix adhesion [34] and structural remodeling [35]. Similarly, *PIEZO1* displayed high intensity in controls and a slightly lower distribution in case subjects (Figure 2A). CVD links are limited to mechanotransduction [36, 37] and pressure regulation [38, 39].

*RUNX2* showed a lower expression in controls, which was absent across most of the case group with some subjects having little or no expression at all. This marker has a crucial role in vascular calcification [40], fibrosis, and arterial stiffness [41] causing inflammation linked to matrix remodeling in CVD progression [42]. The remaining AS markers, *SPP1* and *TGFβ3* did not show any significant variation in expression as it was too low to compare while *BMP2*, *SOX9*, and *AREG* had no expression across the comparison groups. The expression profiling and highest fold enrichment analysis of AS genes revealed Hippo, PI3K-AKT, and TGF-β as statistically significant signaling pathways which play a vital role in the interconnected networks controlling cell growth, survival, and proliferation of cardiomyocytes, differentiation, and inflammation. Crosstalk of these pathways regulates heart development and repair, while their dysregulation contributes to HF, hypertrophy, and vascular disease, making them key therapeutic targets.

#### LVDD-associated gene expression analysis

LVDD-specific gene expression identified heatmap-based expression for eight markers with major characterized by dominant structural and stress-response markers (Figure 2B). *MYH9* and *NDRG1* showed comparably higher expression in the control groups with significant variations observed in case subsets. *MYBPC3* and *BIRC5* showed a moderately lower expression. *MYH9* expressed solid high intensity in control subjects, while expression was significantly variable in case group (Figure 2B). CVD associations include hypertension [43] and cardiac remodeling [44]. *NDRG1* also exhibited consistently high intensity in the control group, while showing considerably variable and lower intensity in the case group (Figure 2B). Its CVD associations involve cellular stress [45] response in myocardial tissues. *MYBPC3* showed moderately low expression in controls in comparison to LVDD cases where the expression dropped further (Figure 2B). CVD associations include HF and hypertrophic cardiomyopathy [46]. *BIRC5* also displayed faintly low expression in the case cohort, slightly more visible in the control group (Figure 2B). It is known to be involved in cell survival signaling in HF [47]. Very low expression was observed in *TGFB2, GDF15, COL3A1, and ANKRD* across both groups (Figure 2B). These genes are associated with syndromic aortic aneurysm, metabolic survival proteins, and cardiac ankyrin repeat protein activity [48–51]. The expression profiling and highest fold enrichment analysis of LVDD genes revealed Hippo signaling, the AGE-RAGE pathway and cytoskeletal dynamics as the statistically significant pathways. The AGE-RAGE signaling significantly contributes to cardiomyopathy and CVD triggering inflammation, and damaging cytoskeletal integrity, particularly altering actin can lead to cell damage, fibrosis, hypertrophy, and impaired heart function. This disrupts cardiomyocyte structure and function with cytoskeletal changes playing a crucial role in cellular stiffness and contractility, promoting conditions like HF.

#### Gene-expression analysis of genes shared between AS and LVDD

The gene expression analysis of the subset of genes shared between LVDD and AS reflected interrelated role and shared molecular biology functions in the diseases (Figure 2C). *TGFB1, TIMP1, and RHOA* expressed very highly in the control group with subset for patients in the case groups showing considerably lower expression. They are critically involved in cardiovascular fibrosis and remodeling [52–55]. *MMP9 and ITGB1* also showed considerably high expression with distinctly lower expression in the case group. They are primarily involved in extracellular matrix (ECM) remodeling, inflammation, and cellular migration [56–59]. *NOTCH1, ROCK1,* and *FOXO1* had low expressions in the case group while little or no expression was observed in the control group. They are linked to chronic hypoxic cardiomyocyte conditions [60–62]. *LGALS3* showed consistently high in both cases and control group. Although it is a robust and active biomarker of chronic inflammation and HF regardless of the specific phenotype it did not any considerable variation between the comparison groups. *MPO, TNF, ACE, NOS3,* and *FAK* showed insignificant expression across the comparison groups. Statistical analysis revealed AGE-RAGE, cardiomyopathy, stress in atherosclerosis, and TGFB signaling as significant with respect to shared genes. The AGE–RAGE axis plays a key role in cardiovascular disease by promoting oxidative stress, inflammation, and endothelial dysfunction, thereby accelerating atherosclerosis and myocardial injury. In cardiomyopathy, chronic metabolic and inflammatory stress activates dysfunctional signaling pathways that impair cardiac structure and function. Cellular stress responses, including oxidative and mechanical stress, further contribute to vascular inflammation and plaque progression in atherosclerosis. Additionally, dysregulated TGF-β signaling is central to cardiac remodeling and fibrosis, influencing vascular stiffness, myocardial hypertrophy, and disease progression across multiple forms of CVD.

## Discussion

### Molecular drivers of AS mineralization

Expression profiling of AS genes identified Hippo, PI3K–AKT, and TGF-β signaling as significantly enriched pathways [63], highlighting their roles in regulating cardiomyocyte growth, survival, differentiation, and inflammation. Crosstalk among these pathways governs cardiac development and repair, while their dysregulation promotes hypertrophy, vascular disease, and HF. Enrichment of these genes was dominated by mechanotransduction and structural remodeling, including Hippo signaling, PI3K–Akt signaling, ECM–receptor interaction, and focal adhesion, underscoring the role of mechanical stress in valvular and early cardiac remodeling. Reduced expression of cell–matrix adhesion and mechanosensory genes in cases suggests impaired cellular responses to pressure overload and progressive disruption of the valvular interstitium. AS progression reflects a transition of valvular cells toward an osteoblast-like phenotype [64]. The low expression of AS genes likely drives the pro-calcific microenvironment and accelerates valvular stiffening as mechanical signaling becomes increasingly dysregulated [65].

### Genomic landscape of LVDD pathogenesis

LVDD genes showed enrichment in Hippo signaling, AGE–RAGE signaling, and cytoskeletal dynamics as dominant pathways [66]. AGE–RAGE activation contributes to inflammation, oxidative stress, and cytoskeletal damage, promoting fibrosis, hypertrophy, and impaired myocardial relaxation [67]. Alterations in actin and sarcomeric components directly affect cardiomyocyte stiffness and contractility, central features of diastolic dysfunction [68]. LVDD-specific genes showed the highest enrichment in cytoskeletal components with significant overrepresentation of hypertrophic, dilated, and diabetic cardiomyopathy pathways, reinforcing the role of structural and metabolic stress [69]. The reduced expression of these genes in cases indicates loss of transcriptional stability in pathways essential for myocardial integrity and mechanical load management. Downregulation of key regulators of contraction and relaxation likely impairs calcium sensitivity, contributing to delayed relaxation and disease progression.

### Shared molecular frameworks for myocardial and valvular remodeling

Shared gene analysis revealed significant enrichment of AGE–RAGE signaling, cardiomyopathy pathways, stress-related atherosclerosis mechanisms, and TGF-β signaling, defining a common molecular response to chronic pressure overload. The AGE–RAGE axis promotes inflammation, oxidative stress, endothelial dysfunction, and matrix remodeling. It alters the actin cytoskeleton modifying the structural integrity of the heart muscles. While dysregulated TGF-β signaling drives fibrosis, myocardial hypertrophy, and tissue stiffening across cardiovascular phenotypes. Gene enrichment further highlighted hypertrophic cardiomyopathy, TGF-β signaling, and the renin–angiotensin system, linking systemic stressors such as hypertension to local myocardial and valvular remodeling. Cardiac remodeling potentially is directed by how the heart adapts its structure to maintain function under different types of stress. High-expression functional analysis emphasized actin cytoskeleton regulation, leukocyte transendothelial migration, and proteoglycan-related pathways, reflecting active structural reorganization and inflammatory signaling.

Across both AS and LVDD, shared genes converge on inflammation and extracellular matrix (ECM) turnover as core drivers of disease. While ECM-regulatory genes showed stable expression in controls, their heterogeneous and reduced expression in cases suggests a dysregulated “fibrotic flux,” marked by imbalance between matrix synthesis and degradation. Collectively, these findings indicate that AS and LVDD are linked by a unified transcriptomic program of chronic inflammation, impaired mechanosensing, and maladaptive tissue remodeling rather than representing isolated disease processes. Biomarkers with strong associations reported for AS have also been linked to multiple non-CVDs. These include chronic disorders such as rheumatoid arthritis [70], multiple sclerosis [71], type 1 diabetes mellitus [72], chronic kidney disease [73], early-onset inflammatory bowel disease [74], systemic lupus erythematosus [75], ankylosing spondylitis [76], schizophrenia [77], ossification of the posterior longitudinal ligament, and atypical femoral fractures [78]. Several rare diseases have also been associated with these genes, including skeletal anomalies [79] (congenital/genetic forms), Loeys–Dietz syndrome type 5, hypophosphatasia, lymphatic malformation 6, and campomelic dysplasia.

Many non-CVD conditions were found to be associated with the targeted genes. For LVDD-associated genes, linked non-CVD conditions include chronic diseases such as type 1 diabetes mellitus [80], diabetic nephropathy [81], and pelvic organ prolapse; rare diseases including vascular Ehlers–Danlos syndrome [82], Loeys–Dietz syndrome type 4 [83], MYH9-related disorder, and Charcot–Marie–Tooth disease type 4D; as well as several cancers, including lung cancer [84], breast cancer [85], pancreatic cancer [86], and adamantinomatous craniopharyngioma [87]. Genes shared between AS and LVDD have also been linked to multiple chronic diseases, cancers, and rare diseases [88]. Detailed classification is provided in Table 2 (Figure 3A and C). In the gene expression analysis, the distribution of age at the time of HF diagnosis across genders indicated a slighter earlier diagnosis in males (45 years and older) compared with females (51 years and older). Similarly, age distribution based on variant profiling revealed an earlier onset of the disease in males (29 years and older) than in females (51 years and older).

## Conclusion

To investigate the genetic basis of the interconnected pathology of AS and LVDD, we performed a multi-omics blood-based analysis of HF patients. Integration of genetic variants and transcriptional profiles with pathway and disease enrichment analyses revealed distinct and overlapping molecular signatures associated with AS and LVDD. Notably, the rate of shared genetic markers was higher than that observed for AS- or LVDD-specific markers, supporting a strong biological relationship between the two conditions. These findings, across age and gender, validated through state-of-the-art literature, gene–disease annotation databases, and electronic health records, highlight the power of blood-based multi-omics profiling to capture early detection markers linking valvular and myocardial dysfunction. As a minimally invasive and clinically scalable approach, peripheral blood analysis provides an important foundation for the development of precision diagnostics and targeted therapies for patients at risk of HFpEF. While tissue-specific next-generation sequencing, may further refine cell-level mechanisms, blood-based genomic signatures offer a practical and translational framework for early risk stratification and personalized disease management.

## Data Availability

Available on request.

## List of Abbreviations

Aortic Stenosis (AS)

Burrows–Wheeler Aligner (BWA) Cardiovascular disease (CVD)

Database of Single Nucleotide Polymorphisms (dbSNP) Extracellular Matrix (ECM)

False Discovery Rate (FDR) Genome Analysis Toolkit (GATK)

Genome Aggregation Database (gnomAD) Gene Ontology (GO)

Heart Failure (HF)

Heart failure with preserved ejection fraction (HFpEF) Human Phenotype Ontology (HPO)

Human Henome 38 (hg38) Left Ventricular (LV)

Left Ventricular Diastolic Dysfunction (LVDD) Institutional Review Board (IRB)

Kyoto Encyclopedia of Genes and Genomes (KEGG) Non-cardiovascular disease (non-CVD)

Next-generation sequence (NGS)

Peripheral blood mononuclear cells (PBMCs) Reactome Knowledgebase

RNA by Expectation Maximization (RSEM)

RNA Sequencing (RNA-seq) Variant Call Format (VCF) Variant Effect Predictor (VEP) Whole genome sequence (WGS)

## Acknowledgments

This research was conducted with the great support of Division of Cardiovascular Diseases and Hypertension, Department of Medicine, Robert Wood Johnson Medical School (RWJMS), and Rutgers Institute for Health, Health Care Policy, and Aging Research (IFH), Rutgers Health, NJ.

We express sincere gratitude to the current and former members of Ahmed Lab at Rutgers RWJMS/IFH, and appreciate all colleagues, collaborators and institutions who provided direct and indirect insight and expertise that greatly assisted the research and development of this project.

We acknowledge the Office of Advanced Research Computing (OARC) at Rutgers University for providing access to the Amarel cluster and associated research computing resources that have contributed to the development and testing of this application.

## Author contributions

Z.A. led this study; participated in patient consenting, sample collection, and management; generated WGS and RNA-seq data; implemented genomic pipelines to process sequence data; computed gene expressions and discovered variants. P.G. performed bioinformatics analysis to extract, annotate, and visualize target variants. J.M. performed bioinformatics analysis to analyze gene expression and enrichment, and visualized results in heatmaps and bubble plots. N.Y. and P.S. proposed and guided this study and provided strong collaborative support. Z.A. drafted and edited manuscript and figures, and all authors participated in writing, reviewed and approved this study for publication

## Biographical Note

Z.A. is an Assistant Professor at the Department of Medicine, Division of Cardiovascular Diseases and Hypertension, Robert Wood Johnson Medical School (RWJMS), and a Core Faculty Member at the Rutgers Institute for Health, Health Care Policy, and Aging Research (IFH), Rutgers Health. Furthermore, ZA is the Adjunct Assistant Professor at the School of Medicine, UConn Health, CT.

P.G. and J.M. are Research Assistant at the Ahmed lab, Rutgers IFH/RWJMS.

N.Y. is an Associate Professor of Medicine, Section Chief of Clinical Research & AI Innovation, and Director of Data Science and Machine Learning Research in the Division of Cardiology at Rutgers Robert Wood Johnson Medical School in New Brunswick, NJ. Additionally, she directs the Center for Innovation at RWJUH, serves as an Editorial Board Member for the American Society of Echocardiography (ASE), and holds roles as Associate Editor for Translational Systems Biology and In Silico Trials.

P.S. is the Henry Rutgers Professor of Cardiology and the Chief of the Division of Cardiovascular Diseases & Hypertension at Rutgers Robert Wood Johnson Medical School and the Chief of Cardiac Services at the Robert Wood Johnson University Hospital.

## Declarations

Ethical Approval and Consent to participate: Informed consent was obtained from all subjects. All human samples were used in accordance with relevant guidelines and regulations, and all experimental protocols were approved by the Institutional Review Board (IRB) at UConn Health and Rutgers University. All procedures performed in studies involving human participants were in accordance with the ethical standards of the institution and the 1964 Helsinki Declaration and its later amendments or comparable ethical standards.

## Consent for publication

Not applicable

## Availability of data and material

Available on request.

## Competing interests

The Authors declare no Competing Financial or Non-Financial Interests.

## Funding

No funding was received.

## Notes

### Competing Interest Statement

The authors have declared no competing interest.

### Author Declarations

Institutional Review Board at Rutgers University.

